# Long-telomeropathy is associated with tumor predisposition syndrome

**DOI:** 10.1101/2024.11.26.24318007

**Authors:** Patrick Allaire, John Mayer, Luke Moat, Rachel Gabor, Jerry W. Shay, Jing He, Chenjie Zeng, Lisa Bastarache, Scott Hebbring

## Abstract

Telomeres protect chromosomal integrity, and telomere length (TL) is influenced by environmental and genetic factors. While short-telomeres are linked to rare telomeropathies, this study explored the hypothesis that a “long-telomeropathy” is associated with a cancer-predisposing syndrome. Using genomic and health data from 113,861 individuals, a trans-ancestry polygenic risk score for TL (PRS_TL_) was developed. A phenome-wide association study (PheWAS) identified 65 tumor traits linked to elevated PRS_TL_. Using this result, a trans-ancestry phenotype risk score for a long-TL (PheRS_LTL_) was develop and validated. Rare variant analyses revealed 13 genes associated with PheRS_LTL_. Individuals who were carriers of these rare variants had a predisposition for long-TL validating original hypothesis. Most of these genes were new to both cancer and telomere biology. In conclusion, this study identified a novel tumor-predisposing syndrome shaped by both common and rare genetic variants, broadening the understanding of telomeropathies to those with a predisposition for long telomeres.

## Introduction

Telomeres are repetitive DNA elements maintained by a variety of proteins that cap chromosomes and function to prevent chromosomal deterioration. Telomeres have been considered a “biological clock” for aging because they shorten during cell division, leading to a gradual loss of telomere length (TL) across the lifespan ^1^. Numerous genes play a critical role in telomere maintenance and regulation, including *TERT* and *TERC*, which encode the holoenzyme telomerase. Maintaining telomere length is essential for life. Cells accumulating exceedingly short telomeres enter cellular senescence and/or apoptosis ^2^. This mechanism is often circumvented during tumorigenesis via telomerase reactivation through TERT’s re-expression or an alternative lengthening of the telomeres (ALT) pathway ^3^.

Beyond age, TL varies among individuals as TL attrition is affected by various factors including lifestyle, diet, environmental exposures, and genetics. Genetic associations with TL have been observed for both rare and common variants. In the common variant space, multiple genome-wide association studies (GWASs) have identified numerous genetic variants contributing to normal TL variation ^4^. Variation in TL explained by common variants has been reported between 4.35-12.8% ^5–7^. In the rare variant space, variants in telomere maintenance genes like *TERT* and *TERC* have been linked to various Mendelian diseases collectively referred to as telomeropathies. Classic telomeropathies (e.g., dyskeratosis congenita) are characterized by extremely short TL, which causes a variety of symptoms (e.g. lung fibrosis, liver disease, and/or bone marrow failure) ^8^ and are linked with genetic anticipation ^9^. Of importance, little is known regarding rare genetic factors that predispose individuals to long telomeres. This may be driven by a perception that long telomeres reflect healthy living with few negative health consequences.

Although healthy living positively correlates with long-TL, growing evidence indicates the genetics of long-TL also has negative health consequences. In 2014, Robles-Espinoza et al. ^10^ and Shi et al. ^11^ mapped the first rare high penetrant melanoma risk variants to *POT1*. Individuals with these variants also exhibited unusually long telomeres. Since then, others have identified additional *POT1* variants linked to additional malignancies, including glioma, thyroid, lymphoma, and leukemia ^12,13^. More recently, *POT1* variants have been linked to hematopoiesis syndrome and risk for numerous benign and malignant tumors ^14^. Lastly, phenome-wide association studies (PheWASs) provide additional evidence that TL, and the common variants that contribute to TL, are risk factors for several different benign and malignant tumors ^6,7,15^.

We hypothesized that genetic variation contributing to long-TL is associated with a unique syndrome distinguishable from classical telomeropathies. We refer to this TL trait as a “long-telomeropathy.” To test this hypothesis, we studied telomere genetics in large cohorts linked to genomics and electronic health record (EHR) data. Results suggest that both common and rare variants contribute to a long-telomeropathy that impacts the risk of a tumor-predisposition syndrome.

## Methods

### Study population

We drew on data from three independent biobanks for this research. The *All of Us* Research Program biobank collects Whole Genome Sequence (WGS) and EHR data from individuals in the United States. Our analyses were limited to the Controlled Tier Dataset version C2022Q4R9 released on April 20th, 2023. This included 113,861 trans-ancestry (European, EUR; African, AFR; and admixed race, AMR) adults with a minimum of 5 years of EHR data (mean = 13.7 years, defined as the difference between the first and last date of entry of an International Classification of Disease (ICD code) (**ST1**). We determined phenotype status from EHR data using ICD codes mapped to phecodes (PheWAS Catalogue version 1.2) (^16^ and https://phewascatalog.org/phecodes). Phenotype status was defined using the “rule of two,” meaning individuals with an ICD code on 2 or more unique dates were described as a case for the code ^17^. We also mapped phecodes to the Human Phenotype Ontology (HPO) for the Phenotype Risk Score (PheRS) analysis using a previously published map ^18,19^.

Cohorts from Marshfield Clinic’s PMRP (n=18,406) ^20^ and Vanderbilt University’s biobank (BioVU) (n=52,795) ^21,22^ were used for validation studies (**ST1**). PMRP is a cohort of 98% white non-Hispanics, with 77% claiming German ancestry. Fifty-seven percent of PMRP were female, the mean age of blood draw was 50, and they had, on average, 33 years of EHR data. In BioVU, the mean age of blood draw was 54, 57.9% were female, and had nearly 12 years of EHR data. Institutional Review Boards at Marshfield Clinic and Vanderbilt University approved using PMRP and BioVU for this research, respectively.

For genetic mapping studies in *All of Us*, we restricted our analyses to exome regions. Variants within exon regions of the Gencode v42 basic transcripts were included. Details can be found at https://support.researchallofus.org/hc/en-us/articles/4614687617556-How-the-All-of-Us-Genomic-data-are-organized. Variant annotation was retrieved from *All of Us* variant annotation tables using canonical transcripts. Analyses were limited to variants with a minor allele frequency (MAF) of <1% in the study population and presumed high functional impact variants (i.e., splice, stop-gain, and missense). Due to *All of Us* privacy policies, all variables with a count less than 20 were only analyzed if they could be combined with other variables. ClinVar ^23^ was used to identify Pathogenic (P) and likely pathogenic (LP) variants in *TP53*, *CHEK2*, and *APC*; variants with conflicting interpretations were excluded.

### Telomere length polygenic risk score (PRS_TL_)

To create a polygenic risk score for TL (PRS_TL_), we evaluated the 2 largest GWASs of TL to date derived from UK Biobank ^5^ and TOPMed ^6^. Using these previously reported GWASs, there were 161 (UK Biobank) and 48 (TOPMed) independently associated SNPs known to be associated with mTL readily available in *All of Us* (**ST2**). We derived the **PRS_TL_** by weighing each variant by the published effect size ^24^. (PRS_TL_=∑ (effect size × test allele count]))

### Phenotype risk score for long-telomeropathy (PheRS_LTL_)

To identify phenotypes associated with PRS_TL_, and to develop a feature set for a long-telomeropathy PheRS (PheRS_LTL_), we utilized similar strategies that would be applied to PRS development. Whereas a PRS is typically derived from GWAS, the PheRS_LTL_ was derived from PheWAS. In this instance, we conducted a trans-ancestry PheWAS on PRS_TL_. We limited PheRS_LTL_ development to tumor trait features associated with long-TL (P<0.05, 65 tumor traits). Like “LD-pruning” in PRS development, all pairwise correlations between tumor phenotypes were measured. If there were 2 or more phenotypes correlated (r^2^>0.2) (**SF1**), the phenotype with the strongest P-value was chosen. Pruning reduced the list to 38 tumor phenotypes.

Again, like PRS development, “P-value thresholding” was applied to quantify a P-value where PRS_TL_ explained most of the variance in PheRS_LTL_. Each tumor phenotype was incrementally added to the PheRS_LTL_ and associated with PRS_TL_, starting with the most significant phenotype. Each phenotype was weighted by the log inverse prevalence of the feature consistent with PheRS methodology ^18,19^. To validate our novel PheRS_LTL_ in an independent population, we repeated the P-value thresholding process in PMRP. The final PheRS_LTL_ consisted of 26 tumor phenotypes.

### Measured Telomere (mTL)

As previously published, measured TL (mTL) from lymphocyte DNA was quantified in PMRP and BioVU via a high throughput quantitative PCR assay ^7^. mTL was estimated on a subset of *All of Us* participants from WGS data derived from lymphocyte DNA using Telomerecat ^25^. Telomerecat was selected over other TL algorithms, given its computational efficiency and ability to analyze cram files. All samples were run with default parameters and a batch correction during the TL estimation step. mTL was initially estimated in 7 males and 7 females who were randomly selected for each year of age (age 18-90; n=1,024). As expected, age (P=3.22×10^-14^) and PRS_TL_ (P=6.70×10^-4^) were associated with mTL in this *All of Us* population. mTL was also acquired in *All of Us* participants with candidate variants of interest. Individuals with multiple P/LP *TP53* variants were removed as they were likely the result of a somatic mutations. After excluding those with a candidate variant and those with a history of a liquid tumor to avoid additional somatic effects on mTL, 969 of the original 1,024 individuals with mTL were used as a reference.

### Statistical Analyses

All dependent variables considered disease-related and continuous (i.e., PheRS_LTL,_ all other PheRSs, and mTL) were associated with TL genetics using a generalized linear model. All binary dependent variables (i.e., phecodes for PheWAS) were associated with TL genetics using logistic regression. FIRTH was applied for rare variant analysis (i.e., ExWAS). Rare variants were limited to those with allele frequency <1% but included only those with an allele count ≥20 based on *All of Us* privacy policies. Gene-based testing was conducted using SKAT ^26^. Sex at birth, the last age documented in EHRs, the first 16 principal components, and EHR length (i.e. age at last visit minus age at first visit) were used as covariates. When mTL was the dependent variable, linear regression was applied using age of enrollment and sex as covariates. The statistical models for each of the analyses are further summarized in **ST3**.

## Results

### Common telomere genetics are associated with cancer syndromes

We initially sought to evaluate if common SNPs that impact TL could influence risk for phenotypes associated with a classic telomeropathy (i.e., dyskeratosis congenita) using pre-existing data from 2 independent biobanks (i.e., PMRP and BioVU). This work was initially done using a PRS_TL_ with SNPs and weights defined by the largest GWAS of mTL conducted in the UK Biobank ^5^. The association between genetically predicted TL (PRS_TL_) and PheRS for dyskeratosis congenita was non-significant (beta=0.01, P=0.11). Given that many other syndromes already have a PheRS defined ^17,18^, and TL is known to be associated with a broad spectrum of diseases ^6,7,15^, we conducted a scan for associations with PRS_TL_ and 384 Mendelian diseases. Interestingly, PRS_TL_ was associated in both PMRP and BioVU with PheRSs for Li-Fraumeni (MIM: 151623; meta-analysis P=2.10×10^-7^, beta=0.103) and familial adenomatous polyposis (FAP) syndrome (MIM:175100; meta P=1.03×10^-10^, beta=0.223). These associations passed a conservative Bonferroni threshold (384 tests, P<1.3×10-^4^) (**SF2.A**). In both instances, individuals with a genetic predisposition for long-TL had an increased risk for these cancer syndromes. Associations for both Li-Fraumeni and FAP were validated in *All of Us* (P= 1.32×10^-17^ and P=4.27×10^-22^, respectively) (**SF2.B**). Given these results, we hypothesized that PRS_TL_ may contribute to a cancer-predisposing syndrome that phenocopied FAP and Li-Fraumeni. Furthermore, we hypothesized that this syndrome was associated with long telomeres. We developed a novel PheRS for long-telomeropathy (PheRS_LTL_) to evaluate this hypothesis.

### Common telomere genetics contribute to a long-telomeropathy phenotype

We conducted a trans-ancestry PheWAS using 2 PRS_TL_ in *All of Us* to identify phenotypes associated with TL genetics. The PRS_TL_ was derived from the 2 largest previously published GWAS of mTL. The first was a GWAS by Codd et al. from the UK Biobank, as mentioned above ^5^. The second was from Taub et al., a smaller but trans-ancestry GWAS of mTL from TOPMed ^6^. As expected, and consistent with previous reports by us and others, ^5,6^ statistically significant phenotypes associated with either PRS_TL_ were mainly tumor-related with increased risk correlating with a genetic predisposition for long telomeres (**Figure 1 and ST5**). When comparing PheWAS results from the 2 PRS_TL_, UK Biobank-PRS_TL_ resulted in fewer significant associations in *All of Us*’s EUR ancestry population (n=19) compared to the TOPMed-PRS_TL_ (n=21). Interestingly, many of the associations observed in the UK Biobank-PRS_TL_ did not transcend across the other genetic ancestries. In comparison, TOPMed-PRS_TL_ PheWAS produced more consistent results across the individual genetic ancestries and resulted in more robust statistically significant phenotypes in the trans-ancestry analysis (TOPMed n=28; UK Biobank n=19). Because of this, we used the TOPMed-PRS_TL_ to generate a trans-ancestry PheRS_LTL_.

**Figure 1:**
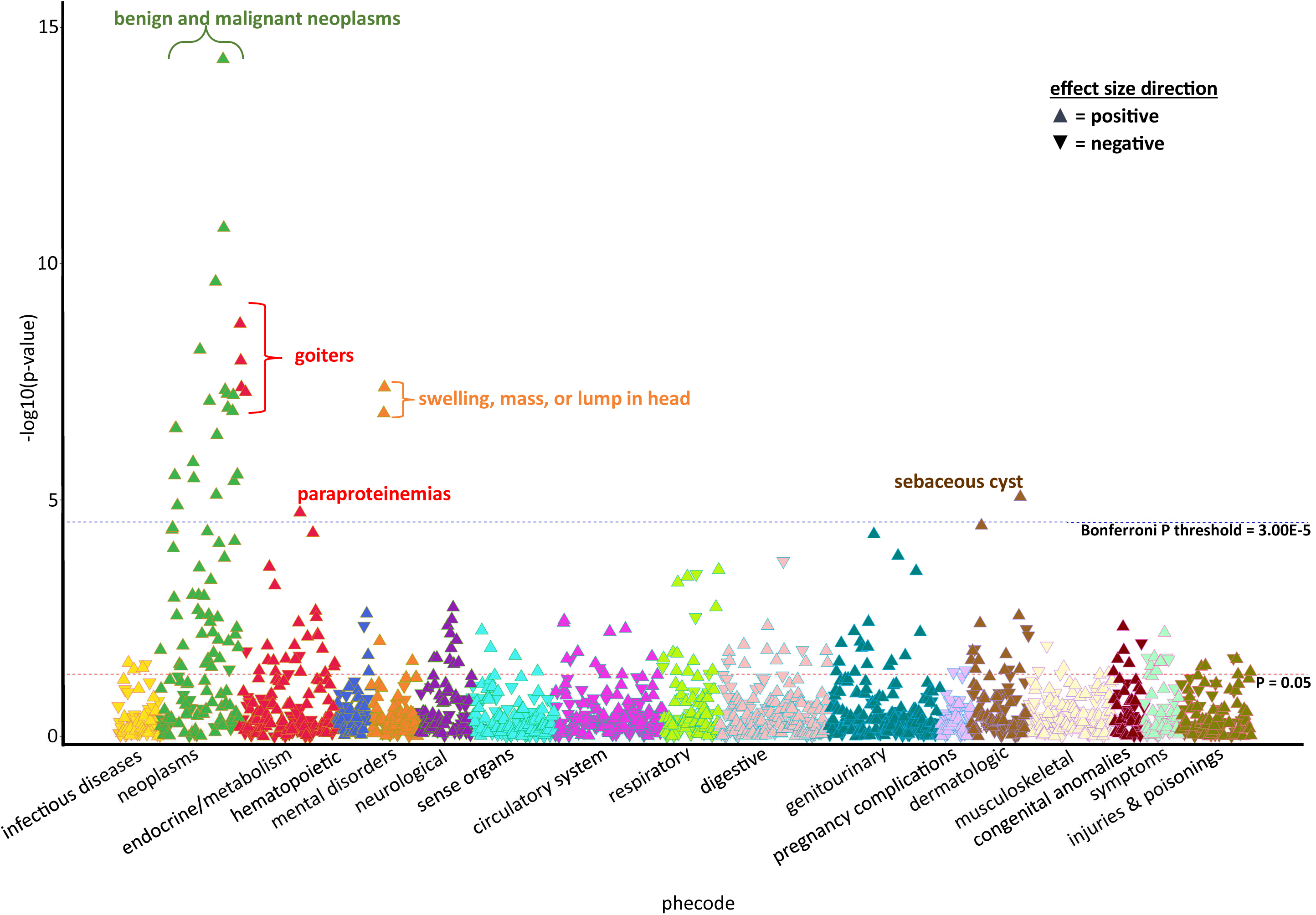
PheWAS of genetically estimated telomere length (TOPMed PRS_TL_). The red lines indicate the threshold for nominal significance, and the blue line represents the threshold for phenome-wide significance.

The PheWAS results for the TOPMed-PRS_TL_ identified 28 statistically significant associations after Bonferonni correction; all were tumor-related, including carcinomas and sarcomas. The strongest associations were lipoma (P=4.20×10^15^), nontoxic nodular goiter (P=1.68×10^9^) and thyroid cancer (P=5.32×10-9). When relaxing the p-value threshold to P<0.05, 65 of 141 tumor traits were associated with PRS_TL_. In total, 26 independent tumor phenotypes were associated with PRS_TL_ after excluding correlated phecodes and p-value thresholding (see methods and **SF1**). The association between PheRS_LTL_ and PRS_TL_ was highly significant (P=5.16×10^-68^) (**SF2**). The PheRS_LTL_ was further validated in the PMRP cohort (**SF3**). Unlike a PRS that weighs SNP by effect size, the PheRS method weights phenotypes according to how common they were in the population ^18,19^. As a result, individuals with low PheRS_LTL_ were enriched for more common benign tumors (**Figure 2A**), whereas those with high scores were enriched for rarer malignant tumors (**Figure 2B**). At opposite extremes of the PheRS_LTL_ distributions, 59% of the population scored 0-1 with a mean z-scored PRS_TL_ of -0.0247. Nearly 2.4% of the population had a PheRS_LTL_ >6 with a mean of 0.1359, a 6.5-fold higher PRS_TL_ than those with a PheRS_LTL_ of 0-1 (**Figure 3A**). Although PRS_TL_ was correlated with PheRS_LTL_ in *All of Us* (r^2^=2.91×10^-3^), this correlation was impacted by age (**Figure 3B/C**). For every decade of adult life, there was an incremental increase in average score and strength in correlation. At ages 18-29, there was a non-significant correlation between PRS_TL_ and PheRS_LTL_ (P=0.218, r^2^=2.79×10^-4^). The strongest correlation between PRS_TL_ and PheRS_LTL_ was for individuals >80 years of age (P=2.25×10^-12^; r^2^=5.52×10^-3^).

**Figure 2:**
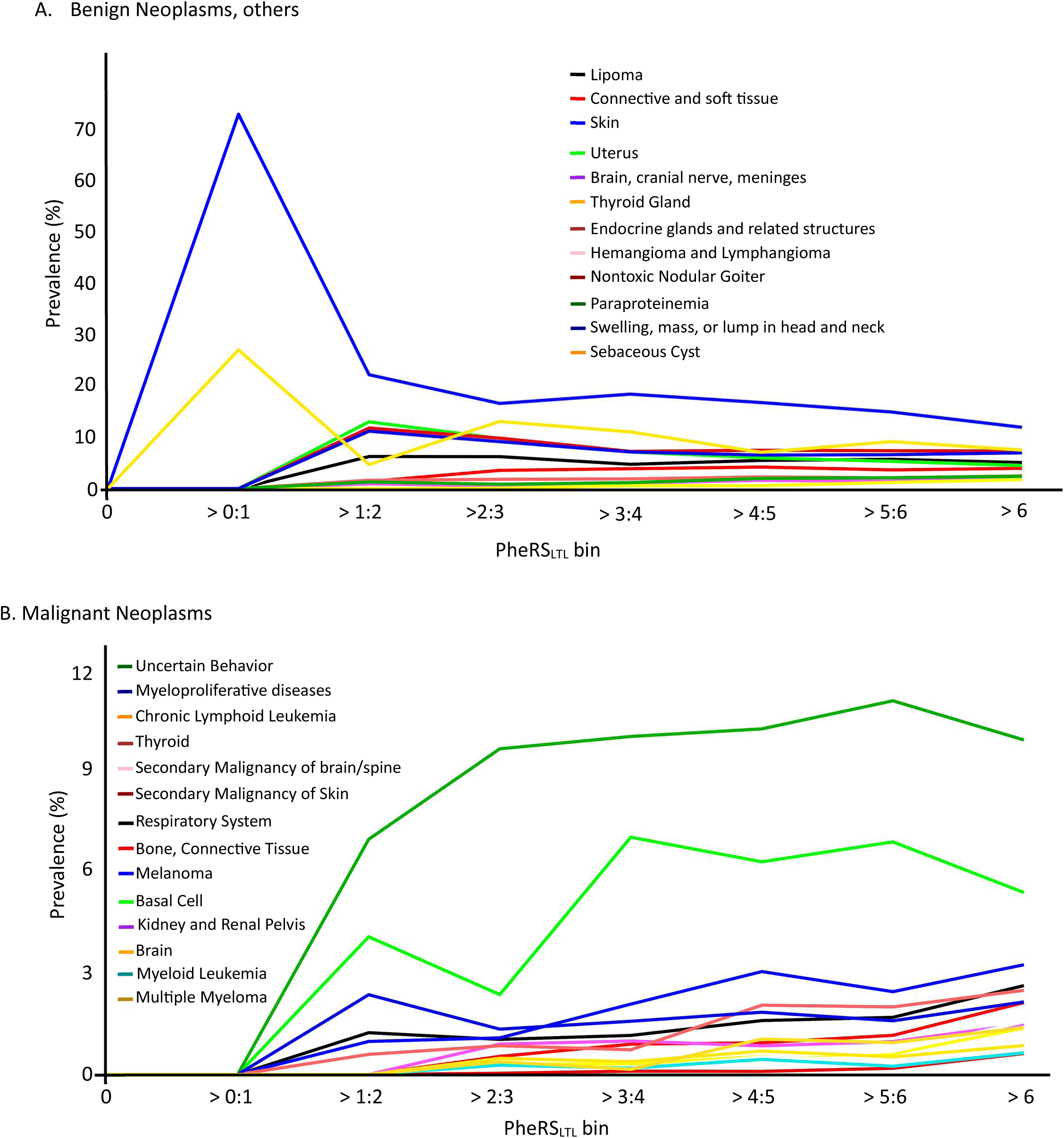
Prevalence of Tumor Cases in the Population Binned by Phenotype Risk Score for Long TL (PheRS_LTL_) **A:** Benign tumors. **B:** Malignant tumors.

**Figure 3:**
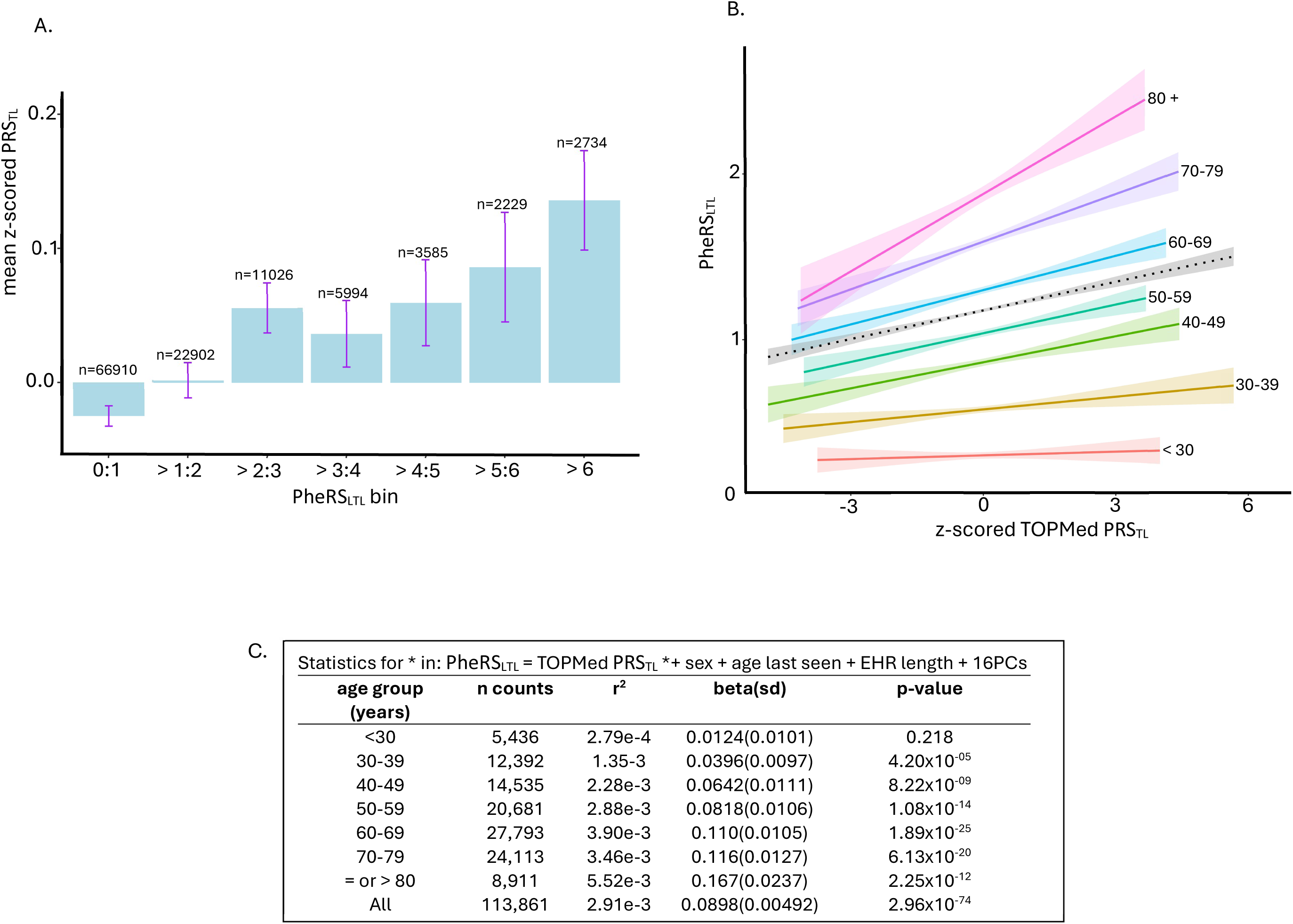
Correlation Between Genetically (TOPMed PRS_TL_) and Phenotypically (PheRS_LTL_) Predicted Telomere Length. **A:** Average PRS_TL_ by PheRS_LTL_ bins, error bars represent 95% confidence intervals. **B:** Linear regression model of PheRS_LTL_ vs PRS_TL_ by age group, shading captures 95% confidence intervals. **C:** Summary statistics derived from the association test in panel B between PheRS_LTL_ and PRS_TL_ by age group.

As mentioned previously, PRS_TL_ was associated with PheRS for Li-Fraumeni and FAP (**SF2**). Variants often influence FAP in *APC,* whereas Li-Fraumeni can be influenced by variants in *TP53* and, to a lesser extent, *CHEK2*. Assuming these known cancer syndromes are primarily independent of telomere genetics, we stratified the population between those with and without rare P/LP variants in *TP53* and *CHEK2* (Li-Fraumeni) and *APC* (FAP) (**ST7**). As expected, those with P/LP variants had higher PheRS_LTL_ than wildtypes (Wilcoxon rank sum test P=3.40×10^-5^) (**SF4A**). We hoped that stratifying the population may improve the correlations between PheRS_LTL_ and PRS_TL_ by removing individuals who may artificially inflate PheRS_LTL_, especially at the extremes. Still, the overall correlation between PheRS_LTL_ and PRS_TL_ remained essentially unchanged after excluding individuals with P/LP variants in *APC*, *TP53* and *CHEK2* (**SF4.B/C**). Association between PRS_TL_ and PheRS_LTL_ in individuals with a genetic risk for a known cancer syndrome (FAP and Li-Fraumeni syndrome) was not significant (P=0.159) (**SF4D**).

We assessed the relationship between mTL, PRS_TL_, and PheRS_LTL_ in Marshfield Clinic’s biobank (**ST4**). There were strong associations between mTL and PRS_TL_ (P=2.59×10^-110^, r^2^=0.0259), PRS_TL_ and PheRS_LTL_ (P=1.18×10^-08^, r^2^=0.00165), and mTL and PheRS_LTL_ (P=1.92×10^-11^, r^2^=0.00431). When both PRS_TL_ and mTL were associated together with PheRS_LTL_, mTL and PRS_TL_ remained significant (P=4.36×10^-9^, r^2^=0.00427 and P=2.32×10^-06^, r^2^=0.00117, respectively). This analysis provided evidence that other TL-dependent factors not captured by PRS_TL_ also contributed to PheRS_LTL_. This may include rare variants not included in the PRS_TL_.

### Rare variants contribute to a long-telomeropathy phenotype

We sought to identify rare variants (MAF<1%) that may contribute to risk for a long-telomeropathy as captured by our PheRS_LTL_. PheRS_LTL_ was evaluated using ExWAS and gene-based (SKAT) analyses. In the ExWAS, 9 variants from 9 genes passed an experiment-wise Bonferroni significance threshold (192,285 tests, P<2.6×10^-7^) (**ST8**). No P-value inflation was observed based on the Q-Q plot (λ=0.963) (**Figure 4**). Two of the 9 genes have a clear role in telomere biology, including *ATM* [P=4.27×10^-17^; NM_000051.4:c.6919C>T (p.Leu2307Phe)] ^27,28^ and *DCLRE1B* [P=1.52×10^-12^; NM_001319947.2:c.1150A>T(p.Asn510Tyr)] ^29–31^.

**Figure 4:**
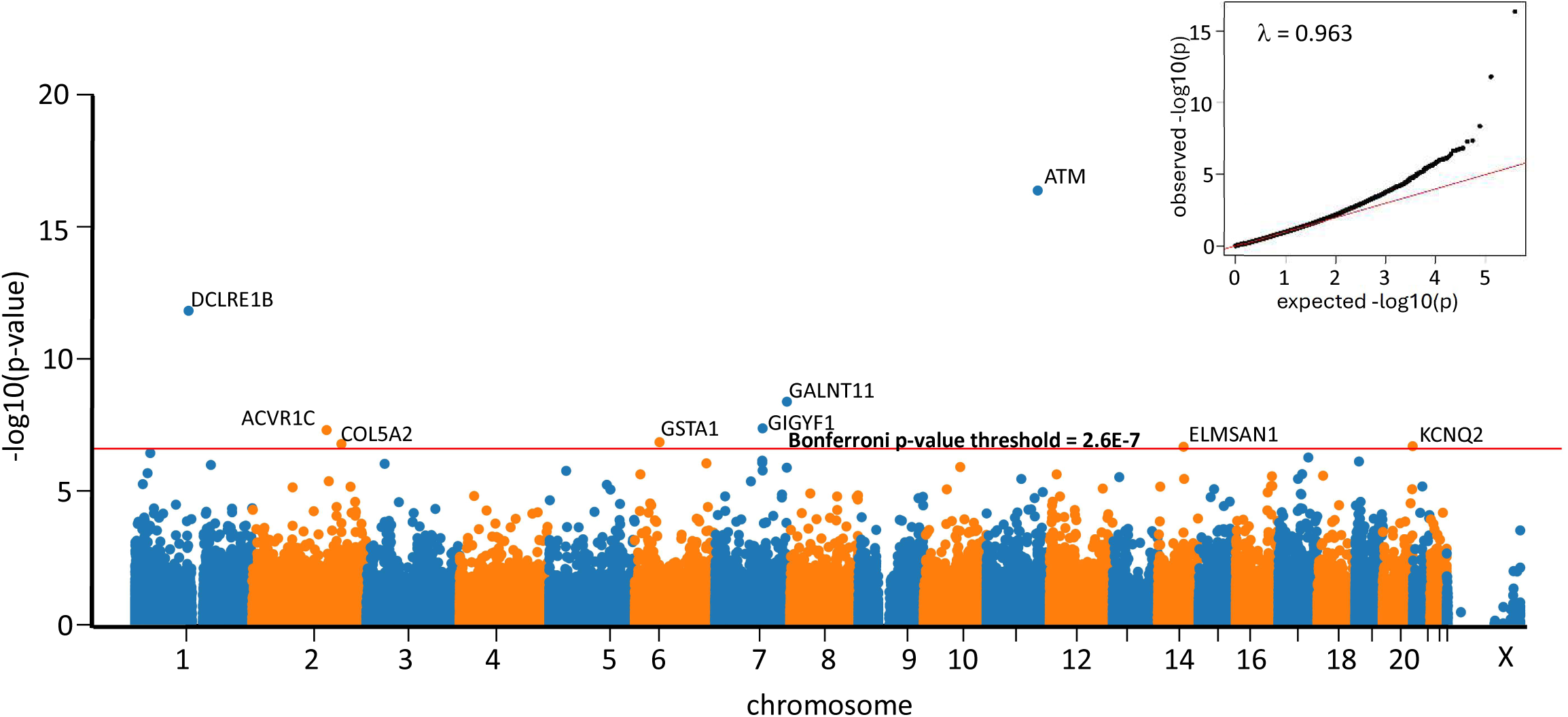
Exome-wide association study (ExWAS) of PheRS_LTL_. **A:** Manhattan plot summarizing p-values. The inset shows the QQ plot. Only variants with protein-altering effects were analyzed. The red line defines the p-value threshold for multiple comparison testing.

SKAT analysis was applied to screen additional associations where multiple rare variants may be associated with PheRS_LTL_ (**ST9**). Five genes were significant (15,746 genes tested, P<3.17×10^-6^), including 2 captured in the ExWAS (i.e., *ATM* and *DCLRE1B*). Just outside the Bonferroni significance threshold was *TERT* (P=3.38×10^-6^), a well-known component of telomerase. No meaningful residual associations were detected via SKAT after removing the top variants (**ST9**). Of relevance, rare P/LP variants in *POT1* are known to result in abnormally long-TL and are risk factors for multiple tumors. Due to allele frequency thresholds, these variants were not captured in the ExWAS or SKAT analysis. As such, it was unsurprising that *POT1* was not associated with PheRS_LTL_ via SKAT (P=0.152).

As a follow-up, and because PheRS_LTL_ is not normally distributed, we performed a permutation test (1,000,000 permutations/variant). The permutation test rejected the null hypothesis for every candidate rare variant, strengthening the original ExWAS and SKAT results (**ST10)**. When evaluating each rare variant individually in relationship to the 26 tumor traits that contribute to PheRS_LTL_, all were associated (P<0.05) with a minimum of at least 2 tumor traits. The *ATM* candidate variant was associated with the most tumor traits (n=13) (**ST11**).

### Rare variants are associated with long telomeres

Given the known relationship between common variants and mTL, along with the relationship between PRS_TL_ and PheRS_LTL_ described above, it was hypothesized that individuals with candidate rare variants would have long telomeres. To test this hypothesis—and to validate the presumed biology driving the associations between PheRS_LTL_ and rare variants—we compared the age, sex, and ancestry-adjusted mTL between individuals carrying at least one allele in the 13 candidate genes compared to a wildtype population in *All of Us*. For a positive control, we identified individuals with one of the *POT1* variants previously linked to long telomeres and similar tumor risks (rs1064794328, rs947005337, and rs587777476) ^14^. We did the same for carriers of P/LP alleles in *APC, CHEK2, and TP53* (**ST7**). Despite a small number of participants with a *POT1* variant (n=15), mTL was found to be, on average, 422 bp longer in *POT1* carriers compared to wild types (422bp, 95% CI: 24bp-821bp, P=0.038). Of the 13 candidate genes, 9 were significantly associated with mTL even after considering multiple comparison testing (P<0.0033). This included *TERT* (135bp, 95% CI: 69bp-201bp, P=5.88×10-5) and *MAP3K3* (176bp, 95% CI: 68bp-283bp, P=0.0010) that were picked up via SKAT but did not reach the statistical significance threshold in the ExWAS. Based on the amplitude of effect, the top genes included *GIGYF1* (324, 95% CI: 141bp-506bp, P=5.1×10^-4^) and *DCLRE1B* (246, 95% CI: 168bp-325bp, P=7.63×10^-10^) (**Figure 5).**

**Figure 5:**
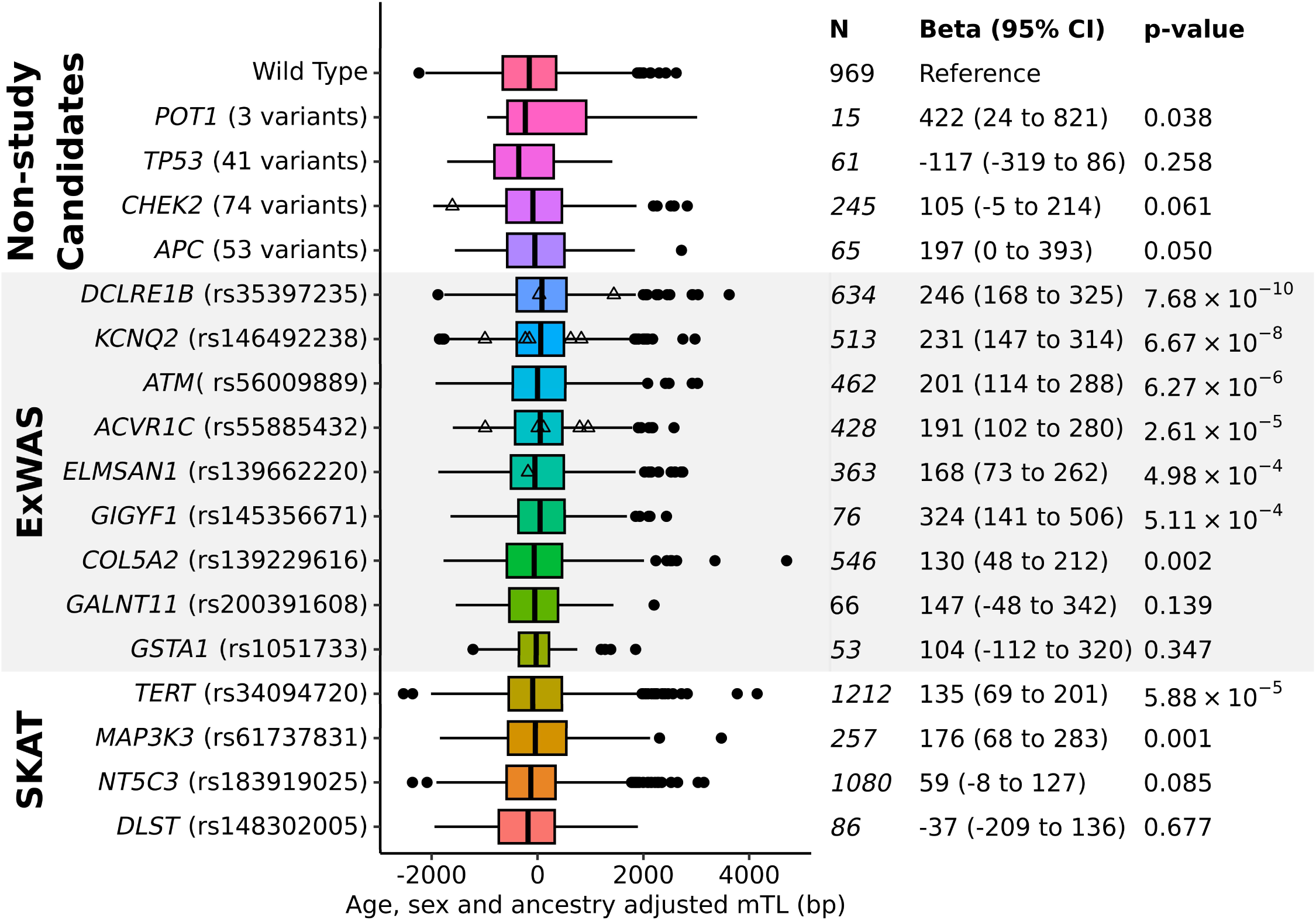
Rare variants are associated with long telomeres. **A.** Density histogram of measured telomere length between wild type and variant carriers. **B.** Statistical analysis of the mean difference between wild type and variant carriers.

### Long-telomeropathy genes have similar effects as known cancer syndrome genes

To better understand the strength of associations between PheRS_LTL_ and TL genetics in the context of other known cancer syndromes (i.e., Li-Fraumeni and FAP), we compared telomere genetics with known P/LP variants (**ST7**) as they related to their corresponding PheRS. In these analyses, the only variables that changed in the linear models were the dependent variables (i.e., PheRS) and their corresponding genetics. All rare variants were evaluated under a dominant model, and the continuous PRS_TL_ variable was binarized based on upper 1, 2, and 3 standard deviations to increase interpretability. The largest effect size of all variants evaluated was between P/LP variants in APC and its corresponding PheRS for FAP (P=1.15×10^-12^, beta=2.02). P/LP variants in *TP53* were also strongly associated with PheRS for Li-Fraumeni syndrome (P=1.54×10^-5^, beta=0.585), but the same could not be said for P/LP variants in *CHEK2* (P=0.814, beta=0.0203). It is well known that *CHEK2* variants often have incomplete penetrance ^32,33^, and its relationship with Li-Fraumeni syndrome remains a topic of debate ^34^. In comparison, candidate variants in *ATM*, *DCLRE1B, GALNT11, GIGYF1, GSTA1,* and *DLST* had comparable or higher effect sizes than P/LP variants in *TP53*. The weakest effect size between telomere genetics and PheRS_LTL_ were common variants defining PRS_TL_ (**Figure 6**).

**Figure 6:**
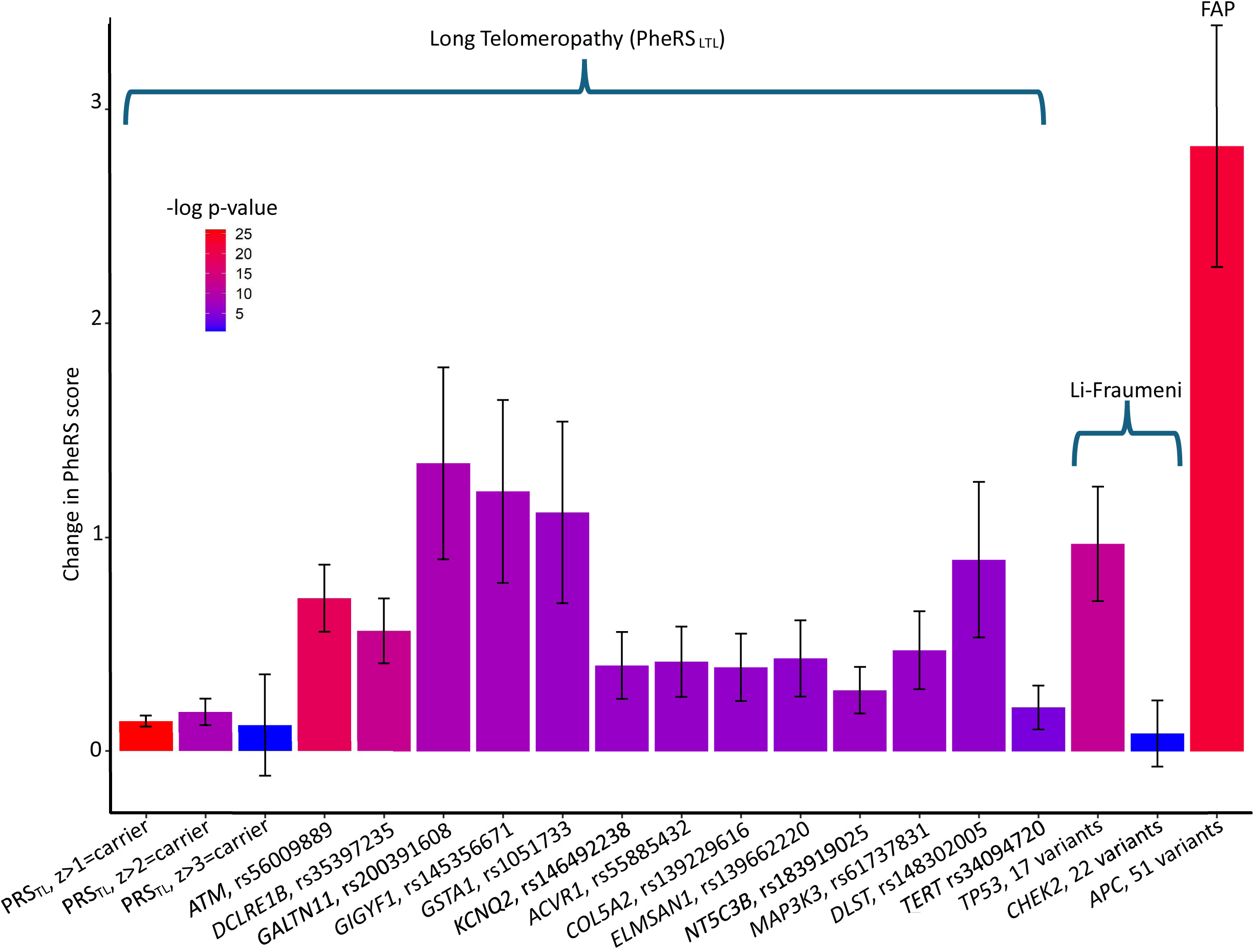
Rare variants contribute to a long-telomeropathy phenotype. Graphed are the effect sizes for genetic variants with their corresponding PheRS. To make all effect sizes comparable, PRS_TL_ was binarized at the upper first, second, or third standard deviation.

## Discussion

This study builds on a growing body of evidence that long telomeres are a significant risk factor for multiple tumor types, including benign and malignant ones. We derived and validated a trans-ancestry PheRS_LTL_ composed of 26 independent tumor phenotypes using common variants previously associated with mTL ^6^ (**ST6**). In nearly all examples, individuals with a genetic predisposition for long telomeres were at risk for carcinomas and sarcomas ranging from malignancies of the skin, brain, bone, lung, thyroid, kidney, and white blood cells (**Figures 1** and **2**). In addition to common variants, we present evidence that rare variants in 13 genes may contribute to a novel and heritable telomere-dependent cancer-predisposing syndrome (**Figure 4** and **ST6** and **ST7**). Some of these variants had effects comparable to P/LP variants linked to other cancer syndromes (**Figure 6**).

Of the 13 candidate rare variants associated with PheRS_LTL_, all but 4 were associated with long mTL (**Figure 5**). Only *DCLRE1B* ^29–31^, *ATM* ^27,28^, and *TERT* ^1,35,36^ are well-known to be associated with telomere length genetics and regulate telomere biology. For the 4 variants not associated with mTL, it is uncertain if the lack of an association was a result of limited power due to small sample sizes, false positives from the original association with PheRS_LTL_, or if these rare variants are still important for tumor risk but via a telomere independent mechanism. Only the variant in *ATM* [NM_000051.4:c.6919C>T (p.Leu2307Phe)] had previously been associated with tumor risk (i.e., lung cancer ^37,38^). More broadly, only 2 of the 13 candidate genes have been implicated in a tumor syndrome (*ATM*, ataxia telangiectasia ^39^ and *DLST*, paraganglioma syndrome ^40^).

The biological mechanism by which long telomeres influence a cancer-predisposing syndrome is not entirely understood. A running hypothesis is that individuals with long telomeres have cells with higher replicative potential than individuals with short telomeres. Therefore, cells with long telomeres can acquire somatic mutations that foster tumorigenesis before telomere-dependent cellular senescence and apoptosis. The precise mechanism of how common and rare variants impact gene function also remains largely unknown. However, some inferences may be made from our study in the context of previous work. For example, DCLRE1B’s exonuclease activity plays a significant role in telomere maintenance by resecting the telomeric DNA. This resection activity facilitates the loading of telomerase onto the telomere, thereby aiding in the elongation of telomeric DNA ^30^. Knockdown of *DCLRE1B* has been reported to induce a DNA damage signal at the telomeres ^29^ and cellular senescence facilitated by the ATM-mediated checkpoint ^41^. Rare loss-of-function variants in *DCLRE1B* are known to cause a form of dyskeratosis congenita and phenotypes consistent of a classical (short) telomeropathy ^31,42^. Because our variant is associated with long telomeres and increased risk for cancer, whereas loss of function *DCLRE1B* variants result in phenotypes consistent with short telomeres, our data infers the p.Asn510Tyr mutation increases DCLRE1B activity. If proven accurate, DCLRE1B may be a candidate for anti-cancer drug development focused on DCLRE1B inhibition.

It is conceivable that PheRS_LTL_ could eventually be used as part of a diagnostic assessment. Individuals with elevated PheRS_LTL_ could be directed to mTL (**Figure 5**) and PRS_TL_ screening (**Figures 3** and **6)**. Even though individuals with elevated PheRS_LTL_ have manifested tumors, they are likely at increased risk for further malignancies where additional cancer screening could be warranted. Although speculative, patients with a long-telomeropathy may also be more responsive to anti-cancer telomerase inhibitors ^3^ than those with average TL. Individuals with a Mendelian form of long-telomeropathy may initiate cascade screening for other family members. Cascade screening may be particularly important for subsequent generations. Genetic anticipation has been reported for those at risk for Mendelian forms of short telomeres (e.g., dyskeratosis congenita), and evidence suggests that the length of telomeres is inherited ^9^. It is plausible genetic anticipation could contribute to long-telomeropathies via the same mechanism.

This study had several limitations. To evaluate clinical applications for PheRS_LTL_ and PRS_TL_, retrospective EHR mining is not likely sufficient. We limited our study population to participants with at least 5 years of EHR data, but all EHR data is influenced by source, demographics, and many other factors ^43^. We show one of those important factors is age, where the relationship between PRS_TL_ and PheRS_LTL_ significantly improved with every decade of life (**Figure 3**). These observations likely reflected an age-dependent penetrance coupled with older individuals having more health history data than younger populations. Conversely, caution is warranted as any focus on older populations may be impacted by survival bias. Regardless, concerns regarding incompleteness and inaccuracies in EHR data, coupled with the age-dependent nature of cancer, would likely inflate only type II errors in our data analyses. Another limitation of our study is restricted to the sample available for this research (i.e, lymphocyte DNA). Nearly all large-scale high-impact TL studies are derived from lymphocyte DNA because blood is a convenient biospecimen. Yet, it is well-recognized that telomere regulation is often tissue-specific ^44^. Because of these inherent qualities, we suspect correlations and effect sizes described throughout our study represent a lower bound for significance. Furthermore, lymphocytes are known to harbor somatic variants indicative of clonal hematopoiesis or a liquid tumor. This is particularly relevant since leukemia was one of the cancer types contributing to PheRS_LTL_. We do not think somatic variants significantly impacted our analyses since some control measures by *All of Us* attempted to filter out such variants ^45,46^. We further removed those diagnosed with a blood tumor during mTL experiments. We also excluded homozygotes/compound heterozygotes for P/LP variants in *TP53,* as these likely arose from somatic mutation. In the end, future family-based studies can be a gold standard for differentiating germline vs somatic variants while providing additional insights into penetrance.

In summary, we show telomeropathies are not limited to just one end of the TL spectrum but should be bifurcated. Whereas short-telomeropathies result in conditions like dyskeratosis congenita, we provide strong evidence that individuals with a long-telomeropathy are at risk for a novel cancer-predisposing syndrome.

## Data Availability

All data produced in the present study are available upon reasonable request to the authors

https://workbench.researchallofus.org/login

## Resource availability

### Lead contact

Dr Scott. Hebbring.

### Material availability

no physical material was used for this manuscript as it is a purely bioinformatic analysis.

### Code and data availability

Only standard data analysis coding (R, Python and SQL) were used. All results and summary dataset are included within the manuscript. Row level data extracted from electronic health record and genomic data from can be retrieved from the All of Us research cohort curated dataset release version 7 (CDRv7) through the workbench and with a “controlled” tiers clearance.

## Acknowledgments

This work was supported in part by NIGMS grants 1R01GM114128 and 1R01GM130715, and the Clinical and Translational Science Award (CTSA) program through the National Center for Advancing Translational Sciences (NCATS), grant UL1TR002373, and generous patients at Marshfield Clinic.

## Author contributions

SH funded the research and provided project oversight. SH and PA wrote the first draft. PA, JM, RG, and JH contributed to data analysis. LM assisted with telomere length experiments. JWS, CZ, and LB were expert consultants. All authors reviewed and approved the final version of the manuscript.

## Declaration of interests

No competing interest to declare

## Supplemental Figure Legends

**Supplemental Figure 1:**
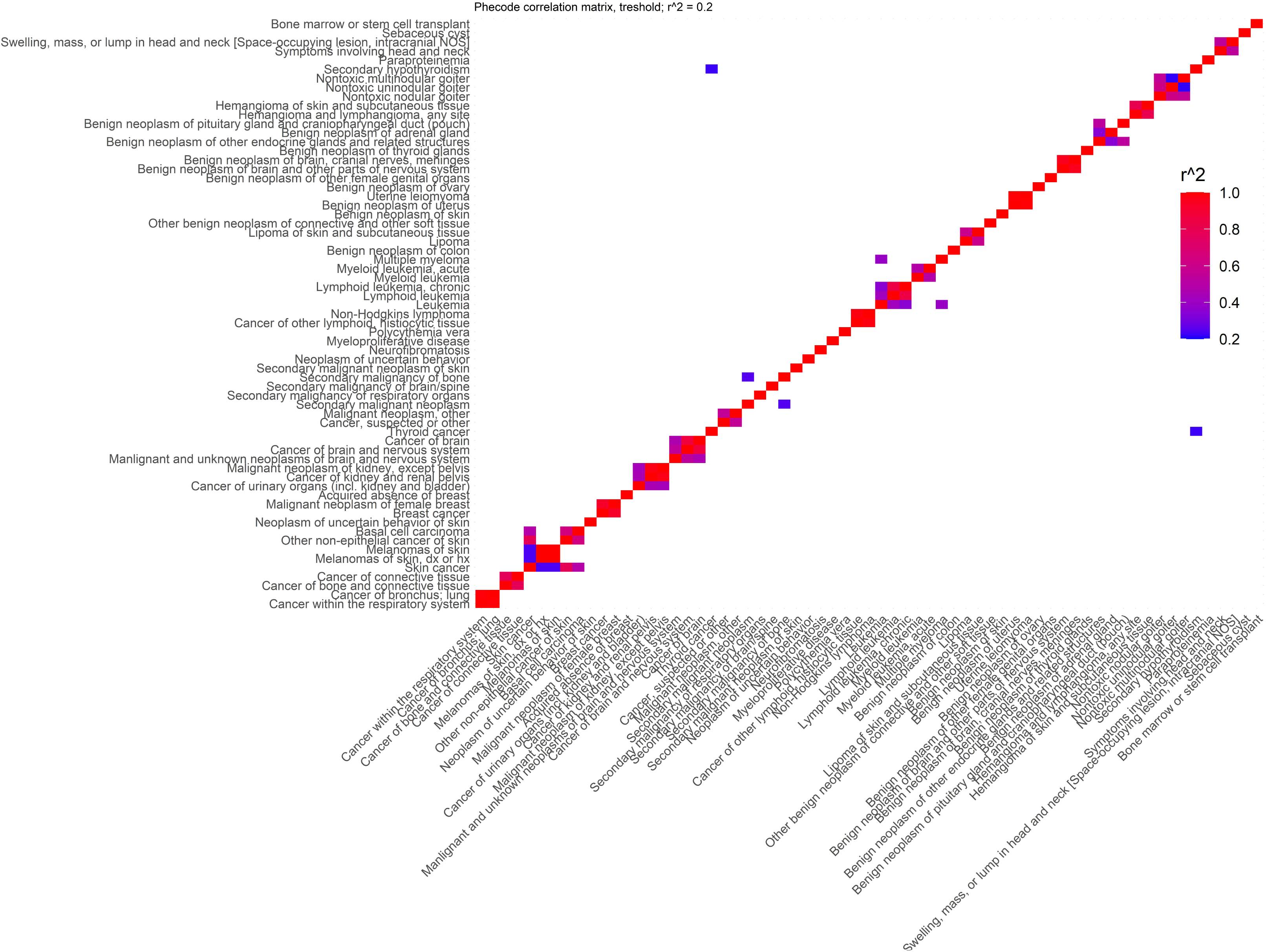
Correlation Matrix of Phenotypes Considered for Establishing PheRSLTL. This includes all phenotypes that passed the Bonferroni adjusted p-value threshold and neoplasms with a p-value < 0.05. For correlating phenotypes with an r^2^ > 0.2, only the phenotype with the lowest p-value was retained for establishing pTL.

**Supplemental Figure 2:**
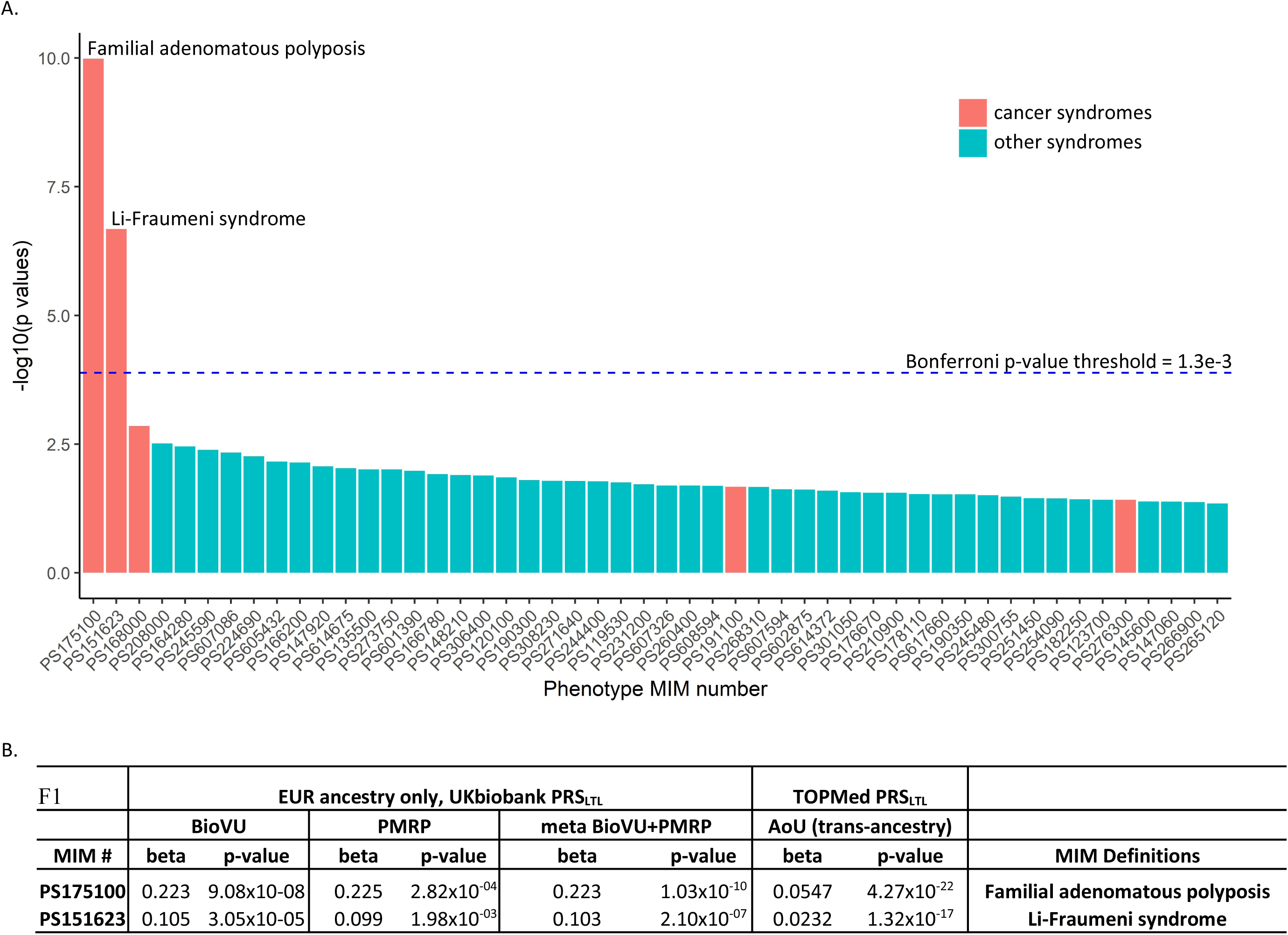
Supplemental Figure 2: MIM PheRS-Wide Association Study of UK Biobank and TOPMed PRSTL. PRSTL was tested for association with the phenotype risk score of various diseases described in OMIM. The model used: PheRS = PRSTL + sex + age last seen + EHR length + first 3 PCs. **Top:** Manhattan plot of screening results with p-values < 0.05. **Bottom:** Summary statistics for results that passed the Bonferroni-adjusted p-value for multiple tests (alpha = 0.05, number of tests = 384, p-value = 1.3e-4).

**Supplemental Figure 3:**
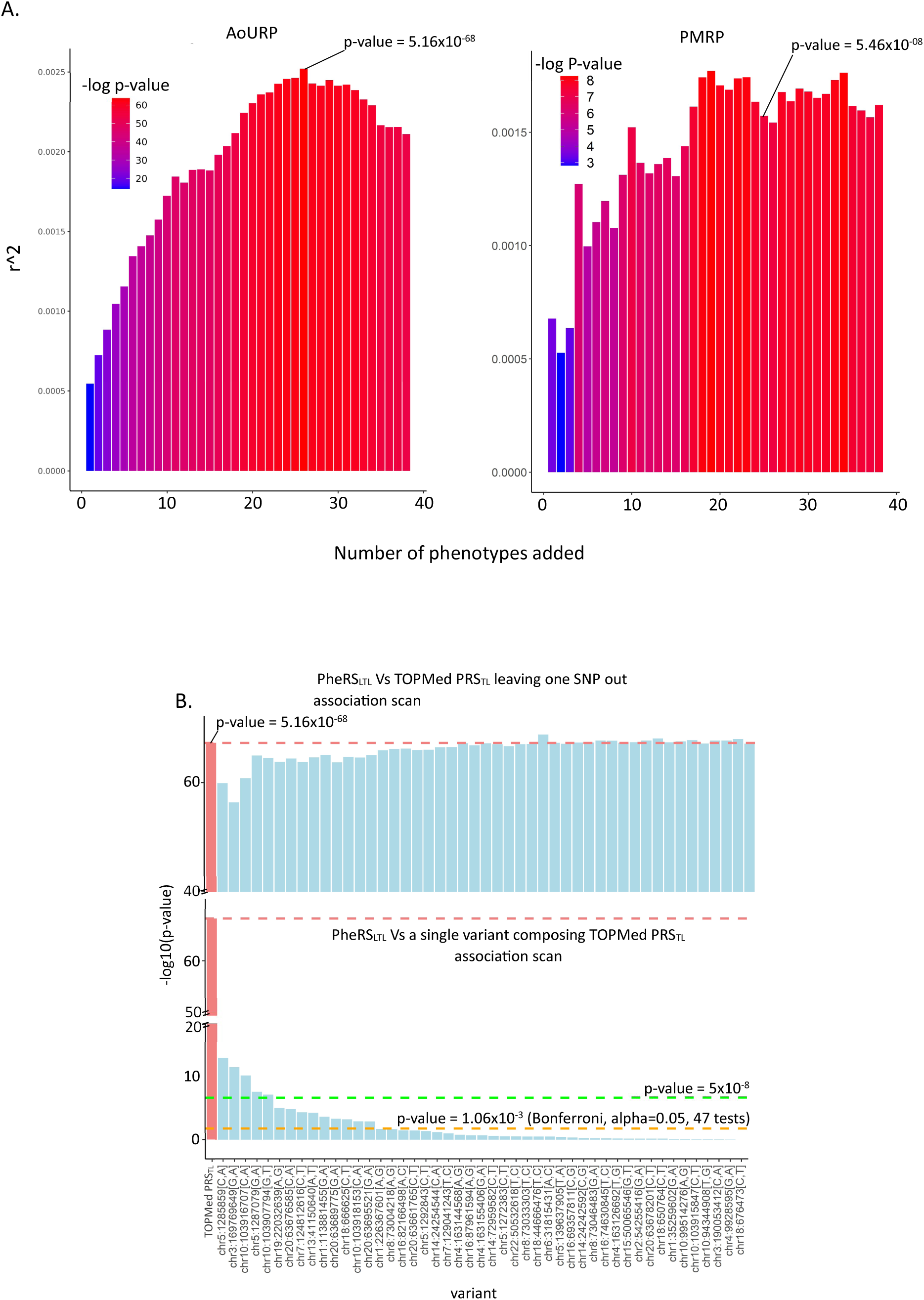
PheRSLTL Selection and Validation. **A:** Selection of phenotypes for inclusion in PheRSLTL and their validation in the PMRP cohort. Individual PheRSLTL was associated with PRSTL using the model PRSTL = PheRSLTL + sex + age last seen + EHR length + first 3 PCs. The statistics for PheRSTL were extracted and plotted to identify the PheRSLTL that best explains the variance in PRSTL. **B:** Examination of the genetic association and contribution of individual SNPs that compose PRSTL to PheRSLTL. **B-Top:** Association between PheRSLTL and PRSTL, excluding one SNP at a time. **B-Bottom:** Association of individual SNPs to PheRSLTL.

**Supplemental Figure 4:**
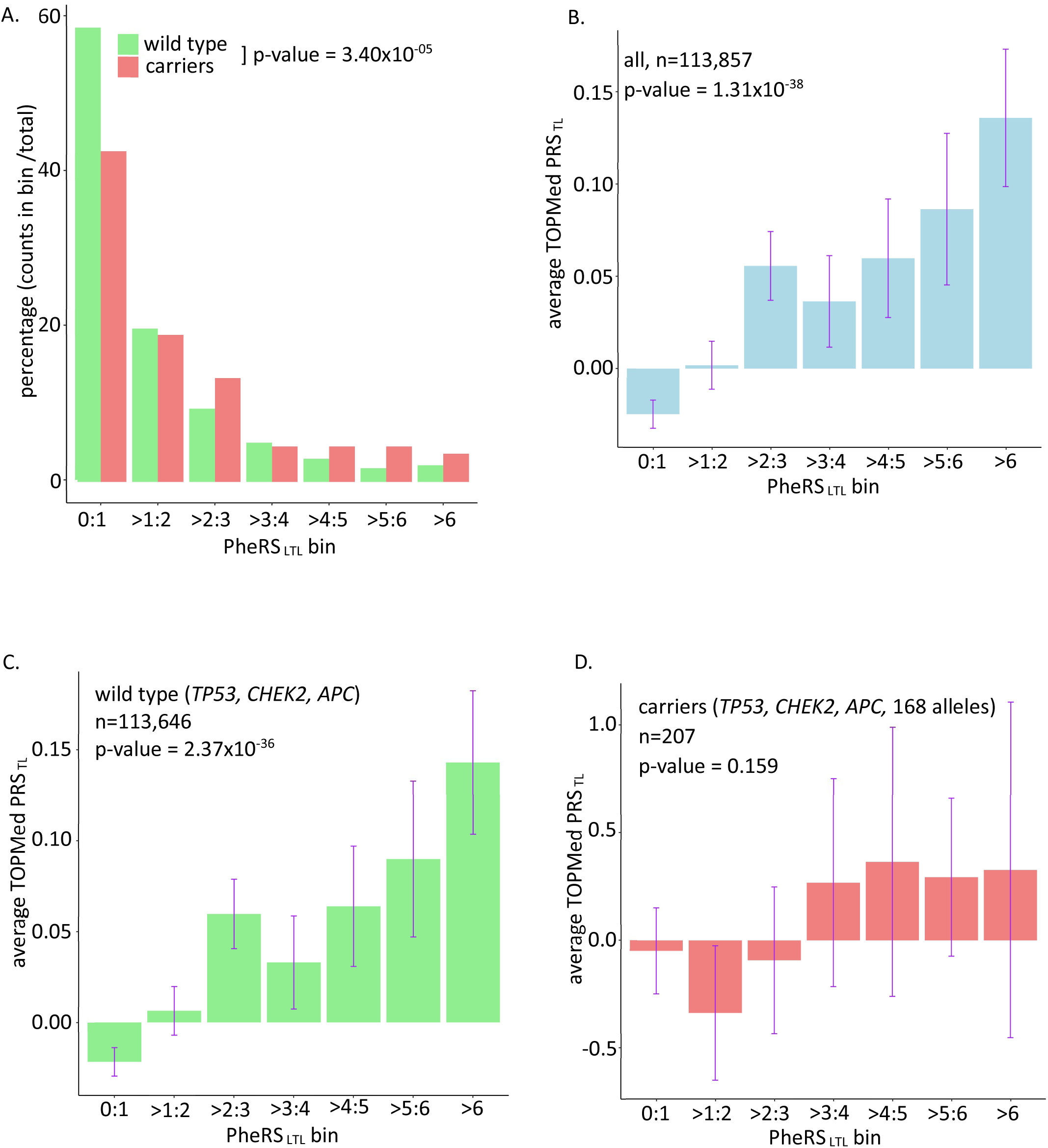
Contribution of *TP53*, *CHEK2*, and *APC* Carriers to the Association of PheRSLTL with PRSTL. **A:** Percentage distribution of wild-type individuals and carriers across PheRSLTL bins. The percentages reflect the total number of wild types and carriers in the cohort. **B:** Average PRSTL per PheRSLTL bin in the entire cohort. **C:** The cohort’s average PRSTL per PheRSLTL bin, excluding carriers. **D:** Average PRSTL per PheRSLTL bin in carriers only. A Wilcoxon rank sum test with continuity correction results in a significance in distribution between raw PheRSLTL wild types and carriers, p-value= 5.46e-06.

## Notes

### Competing Interest Statement

The authors have declared no competing interest.

### Clinical Protocols

https://workbench.researchallofus.org/login

### Author Declarations

Study approved by All of US Institutional Review Board and Marshfield Clinic Research Institute, Institutional Review Board

